# Virulence factors’ contribution to the pathogenicity of *Candida* species: a study based on the clinical isolates of Jaipur, Rajasthan, India

**DOI:** 10.1101/2023.05.06.23289519

**Authors:** Tamanna Pamnani, Rekha Seervi, Parasmani, Blessy Jose, Aakanksha Kalra

## Abstract

In present era, both the Health Care Associated and community acquired infections are increasing steadily. The Leading International Fungal Education (LIFE) has observed that more than ∼80% of world’s population is affected by serious fungal infections. *Candida* species can colonize biotic or abiotic surfaces and transmit via them. *Candida* genus consist more than 200 species with few of infection causing species. *Candida albicans* is one of the species which causes the majority of infections (∼80%) while other species such as *C. glabrata, C. dubliniensis, C. parapsilosis* and *C. krusei* have recently been emerging in these infections. They reside within the body and become pathogenic as the immunity compromised of individual. Virulence factors contribute to spread, transmit and enhance the pathogenicity of the species. The study is focused on analyzing virulence factors of the *Candida* isolates obtained from patients of Jaipur, Rajasthan. Isolates were screened for germ tube formation and production of extracellular enzyme. Results of the study show that along with albican species, non-albicans are also emerging as pathogens. Observations state that blood sample type is most infectious. The study concludes with the statistical analysis of population crucial for preparation of prevention methods and cures to deal this life-threatening disease.

## Introduction

Microbial association with the human body has been known since ages wherein both stay in harmony with each other. *Candida* species is the one such microbe present in the gastro-intestinal tract or vaginal membrane [1].

Usually, *Candida* species can colonize biotic or abiotic surfaces and transmit via them. There are more than 200 members yet only a few (8-10 species) are pathogenic causing Candidiasis. *Candida albicans* is responsible for the majority of infections (∼80%) while other species such as *C. glabrata, C. dubliniensis, C. parapsilosis* and *C. krusei* have recently been emerging [2].

The Leading International Fungal Education (LIFE) has observed that more than ∼80% of the world’s population is affected by serious fungal infections [3]. Virulence factors help a pathogen to invade tissues, secrete enzymes and increase its pathogenicity such as Phenotypic switching (hyphae formation), extracellular enzyme production, secretion of adherence molecules (biofilm formation), Germ tube formation. Therefore, this study is aimed to characterize the two virulence factors (germ tube formation and extracellular enzyme production) and their comparative analysis.

## Materials & Methods

### *Candida* isolates maintenance and identification

38 *Candida* isolates obtained from different specimen types from different individuals were kindly provided by MCRD-CIRD (Microbial Culture Repository Division -Center for Innovation, Research and Development), Dr. B. Lal Clinical Laboratory Pvt. Ltd. The isolates were maintained on YEPD medium (HiMedia Laboratories, Mumbai, India) at 30°C for 16-18 hrs unless otherwise stated and identified by colony morphology and KOH staining. The demographic analysis of the data was carried out using the patients’ details.

### Virulence factor characterization

Two major virulence factors were analyzed in this study.

### Formation of germ tube

It is primarily used to classify the isolates as albicans and non-albicans. However, the ability to form germ tubes has been observed by *C. albicans* and *C. dubliniensis*. For Germ tube formation analysis, isolates were incubated in uninfected human serum for 3 hours at 37°C and evaluated microscopically. 1000 cells were counted in each sample with both germ tube forming and non-forming cells and percentage of the germ tube forming cells were calculated.

### Analysis of Phospholipase Activity

It determines the invasion capacity of the isolate via plate method (phospholipid extraction, preparation of suspension culture and its inoculation followed by formation of precipitation zone) [5].

#### Phospholipid extraction

Eggs were surface sterilized by 70% ethanol and yolk was aseptically separated from albumin. Yolk was centrifuged at 10,000 rpm for 45 mins at 4°C to isolate phospholipid in supernatant.

#### Suspension culture and inoculation

Overnight primary inoculum was prepared in Sabouraud Dextrose broth (SDB) using isolates at 30°C and then adjusted at an optical density (OD) of 0.2 at 600nm ensuring equal cell mass.

#### Media preparation and phospholipase activity

A specific media consisting of Sabouraud Dextrose agar (SDA) with NaCl, CaCl_2_ and 2% egg yolk supernatant was prepared.10μl of the prepared suspension was added on the plates and incubated at 37°C for 4 days to observe the diameter of colony growth and of precipitation zone [5]. The measurement of phospholipase activity is designated as Pz value (“**Pz = colony diameter/colony diameter + zone of precipitation ”)**.

## Results & Discussions

### Demographic analysis

A total of 38 isolates obtained from different specimen samples were used for the study (Supporting Table 1). Age wise analysis (Figure 1A) of the patients showed that though yeast infects all age groups, almost half of infections belonged to older age groups (> 60 years), suggesting the compromised immune system in this age group. Similarly, a gender-based analysis (Figure 1B) suggested a bias of infections in females owing to prevalence of vaginal and urinary tract infections. However, a very low number of isolates from vaginal samples (only 3) and an equal number of isolates from urine samples of males and females (7 each) (Figure 1C) are contra-indicative of the above-mentioned reason. Moreover, results (Figure 1C) show that half of the isolates were obtained from blood samples followed by urine samples with a few from pus and sputum. While a high number from urino-genital infections is expected, such a rise in blood infections is worrisome requiring immediate attention.

**Table 1:** Phospholipase activity of the isolates: Based on phospholipase production (Pz value) all isolates are grouped in five categories as Very high, high, moderate, poor and non-active.

| S.No. | Pz value range | Positive/<br>Negative | Activity | Blood | Urine | Vaginal<br>swab | pus | Sputum | Total |
| --- | --- | --- | --- | --- | --- | --- | --- | --- | --- |
| 1 | Below 0.7 | Positive(++++) | Very High<br>Phospholipase<br>activity | 4 | 5 | 0 | 0 | 1 | 10 |
| 2 | 0.7-0.79 | Positive(+++) | High<br>Phospholipase<br>activity | 5 | 3 | 1 | 0 | 0 | 9 |
| 3 | 0.8-0.89 | Positive (++) | Moderate<br>Phospholipase<br>activity | 4 | 2 | 1 | 0 | 0 | 7 |
| 4 | 0.90-0.99 | Weak positive<br>(+) | Poor<br>Phospholipase<br>activity | 0 | 1 | 0 | 0 | 0 | 1 |
| 5 | 1 and more than 1 | Negative | Non active | 6 | 3 | 1 | 1 | 0 | 11 |
|  | Total | - |  | 19 | 14 | 3 | 1 | 1 |  |

**Figure 1:**
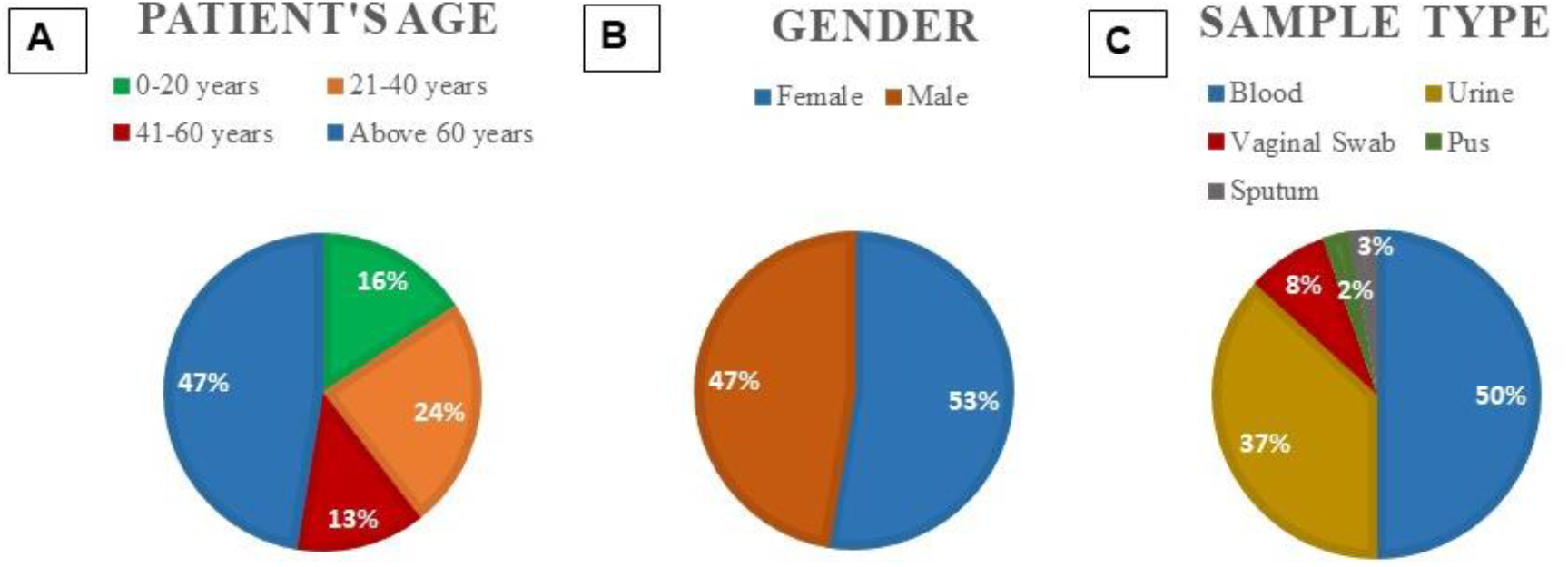
Demographic analysis of isolates: The details of isolates is categorized on the basis of age groups, sex-ration and sample type.

### Analysis of the two major Virulence factors to determine the mechanism of the infections

So far, a total of four major factors have been known to contribute to the virulence out of which, the present study is focused on the characterization of the following factors.

#### i. Formation of germ tube

The representative images of germ tube formation are shown in figure S1. Results (Figure 2A) suggest almost two-third (66%) infections are caused by *albican* or *dubliniensis* species supporting albican to be the major infection causing species. This was also observed with the isolates responsible for causing urino-genital infections (Figure 2C& 2D). However, about one-third infections have been caused by non-albican species suggesting the increase in pathogenic species. It is also observed that more than half of the blood infections have been caused by non-albican species (Figure 2B). The European Confederation of Medical Mycology (ECMM) reported emergence of NAC causing bloodstream infections [6]. It needs to be addressed with utmost priority focusing on identification of antifungal drug molecules, specifically against non-albican species. Also, no such conclusion can be made for isolates from pus and sputum owing to low sample number.

**Figure 2:**
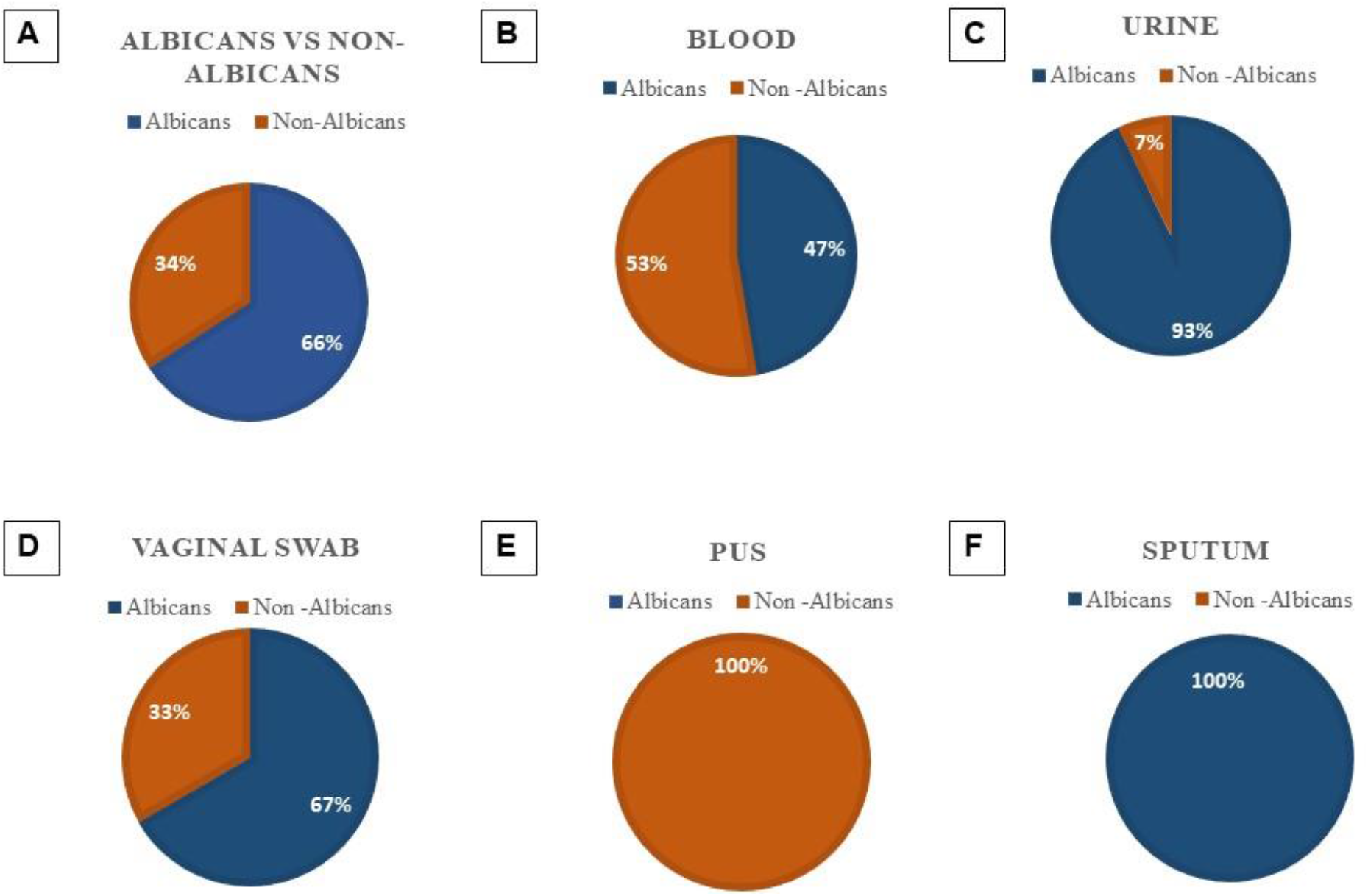
Species distribution based on sample type: The figure shows germ tube formation ability of isolates and distribution based on sample type.

The quantitative analysis suggested that of the 66% isolates producing germ tube, less than 10% germ tube forming cells were found in 84% (21/25) isolates while two isolates each had germ tube producing cells between 10% - 20% and >20% (Table S2). Since these germ tubes are eventually responsible for the adhesion to the host cells [7], this evaluation would be a crucial parameter to assess the severity.

#### ii. Analysis of phospholipase activity

Phospholipase activity is known to play a crucial role in host cell invasion. The calculated Pz values were used to evaluate the phospholipase production wherein the isolates were divided in the five categories: very high (< 0.7), high (0.7 – 0.79), moderate (0.8 – 0.89), poor (0.9 – 0.99) and non-active (1 and above). Representative images of the activity are shown in Figure S2 and the Pz values in Figure3. Results showed that only about 30% isolates were non-active while the rest were almost equally divided among very high, high and moderate categories with a very few isolates possessing poor phospholipase activity (Table 1).

**Figure 3:**
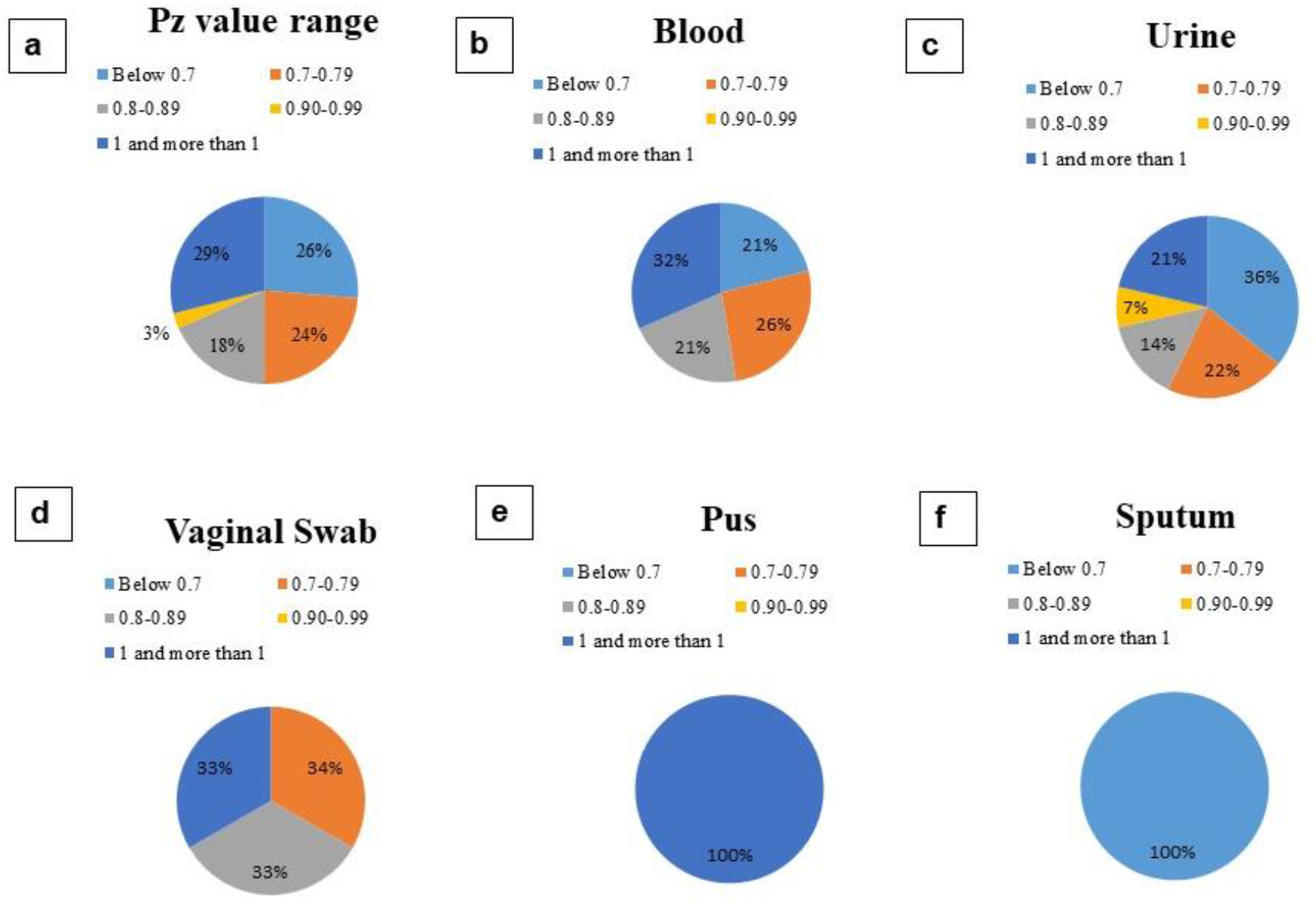
Pz value analysis based on sample type: The figure shows the phospholipase activity (Pz value) distribution on the basis of sample type.

Center for Medical Mycology, Cleveland reviewed and stated that during the infectious phase, *C. albicans* secretes this enzyme, which is crucial for candidal virulence [8]. Although confirmatory research into the phospholipase mechanism(s) is ongoing, preliminary findings suggest direct host cell lysis and damage as the primary process promoting virulence. It was observed that the isolates obtained from urine had significantly high phospholipase producing ability. No phospholipase activity was observed with pus isolate while very high phospholipase activity was observed for the sputum isolate. However, no conclusion can be drawn.

#### iii. Correlation between the two virulence factors

This section of the study was aimed to determine whether any correlation exists between these two activities. Only 12% isolates were non-active for phospholipase in case of germ tube forming cells as against more than 60% in non-germ tube forming ones (Table S3). Not only this, more than 30% of isolates producing germ tube showed very high phospholipase activity as against only 15% in case of non-germ tube producing isolates. It can be concluded that phospholipase activity in each category is significantly higher in the germ tube producers compared to non-germ tube producers. Three plausible explanations can be attributed for this. Firstly, the most virulent isolates possess more than one, if not all, virulence factors at a higher rate. Secondly, germ tube formation is directly or indirectly associated with the expression of extracellular enzymes. Thirdly, non-germ tube forming isolates possess one or both of the other two mechanisms (not evaluated in this study) responsible for virulence of the species. As per analysis at the Department of Microbiology, Italy, it is believed that there is no one element that contributes to the virulence of *Candida*. It is possible that the correlation between phospholipase activity and high GT production in *C. albicans* strains can facilitate mucosal penetration.

Though the germ tube producers generally possess high phospholipase activity, a very low negative correlation (−0.36) was observed between the rate of germ tube formation and level of phospholipase activity (Pz value). Exceptions such as isolate-**12** with very high germ tube formation (11%) but no phospholipase activity or isolate**-1** with 0.3 Pz value but only 1% germ tube forming cells do exist. More mechanistic insights are needed to explain this behavior.

## Conclusion

The recent years have witnessed a significant rise in fungal infections caused by *Candida* both by known species and introduction of novel species. The present study is the first of its kind wherein the isolates obtained from the patients of a tier-2 city of a developing country have been analyzed in terms of virulence factors’ expression. Since *Candida* species are opportunistic, the analysis of the virulence factors is of utmost importance for developing the prevention and treatment strategies. The major conclusions from the study include no age preference or gender bias in infection. The major infections spread in recent years are in blood or in urino-genital tracts. In spite of the emergence of new infection causing species, the majority of infections are still caused by germ tube producers (*albicans* and *dubliniensis*). On a whole, germ tube producing species are more virulent and have better ability to produce phospholipase activity. However, such a direct correlation needs to be validated by further analyses.

## Data Availability

All data produced in the present work are contained in the manuscript.

## Declarations

### Consent to participate

The authors have complete consent for the study.

### Consent for publication

The authors have complete consent for publication.

### Authors’ Contribution

TP performed the experiments and wrote the manuscript; RS and PM performed the experiments; AK designed the research, planned the experiments and wrote the manuscript.

## Acknowledgments

The authors acknowledge Intramural Research Scientific Committee, Dr. B. Lal Institute of Biotechnology, Jaipur, for funding the research.

## Supporting data

The following figures and tables constitute the supporting data for the manuscript.

**Figure S1:**
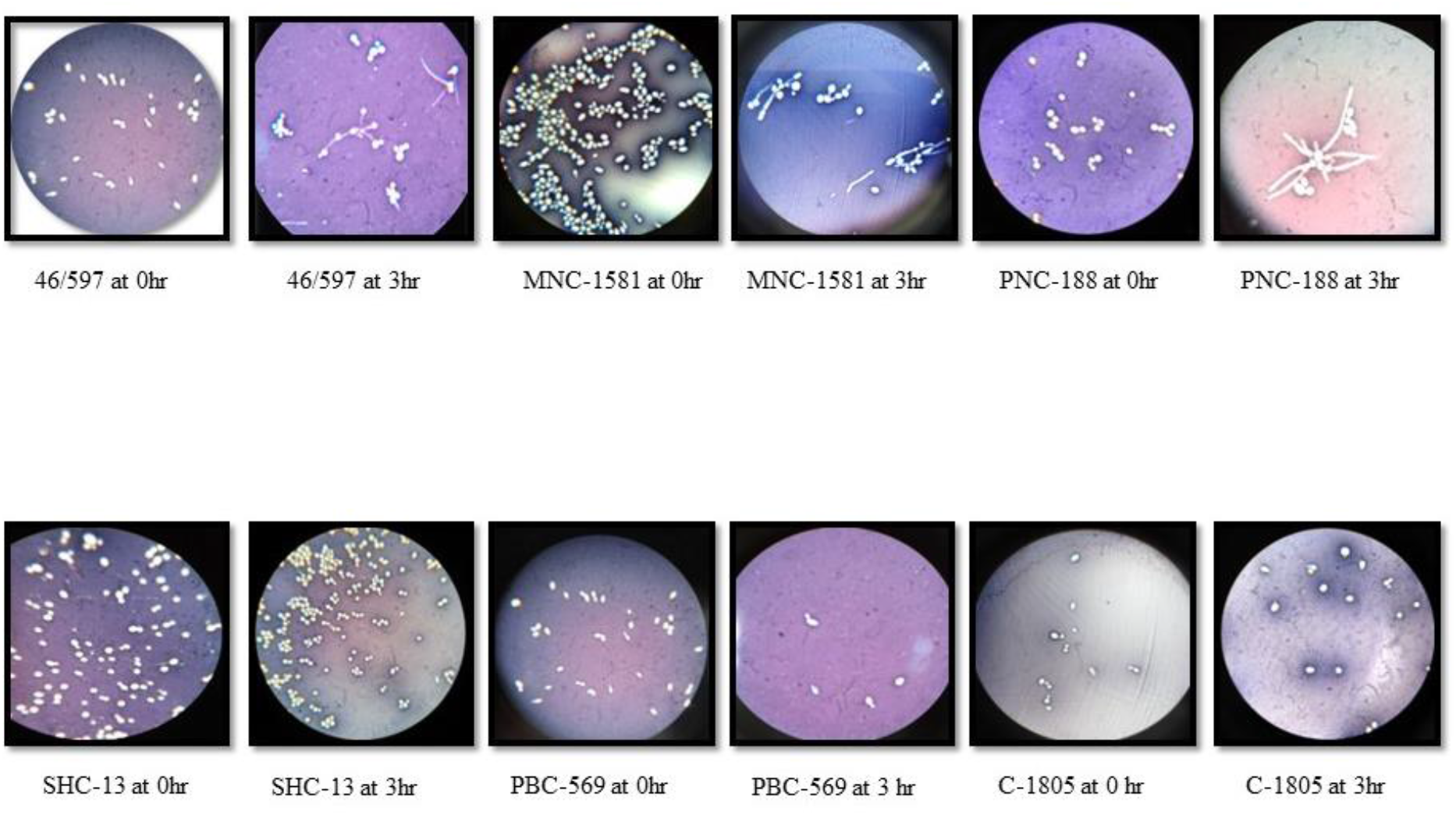
Germ tube formation by the isolates: The figure shows the germ tube formation by the isolates.

**Figure S2:**
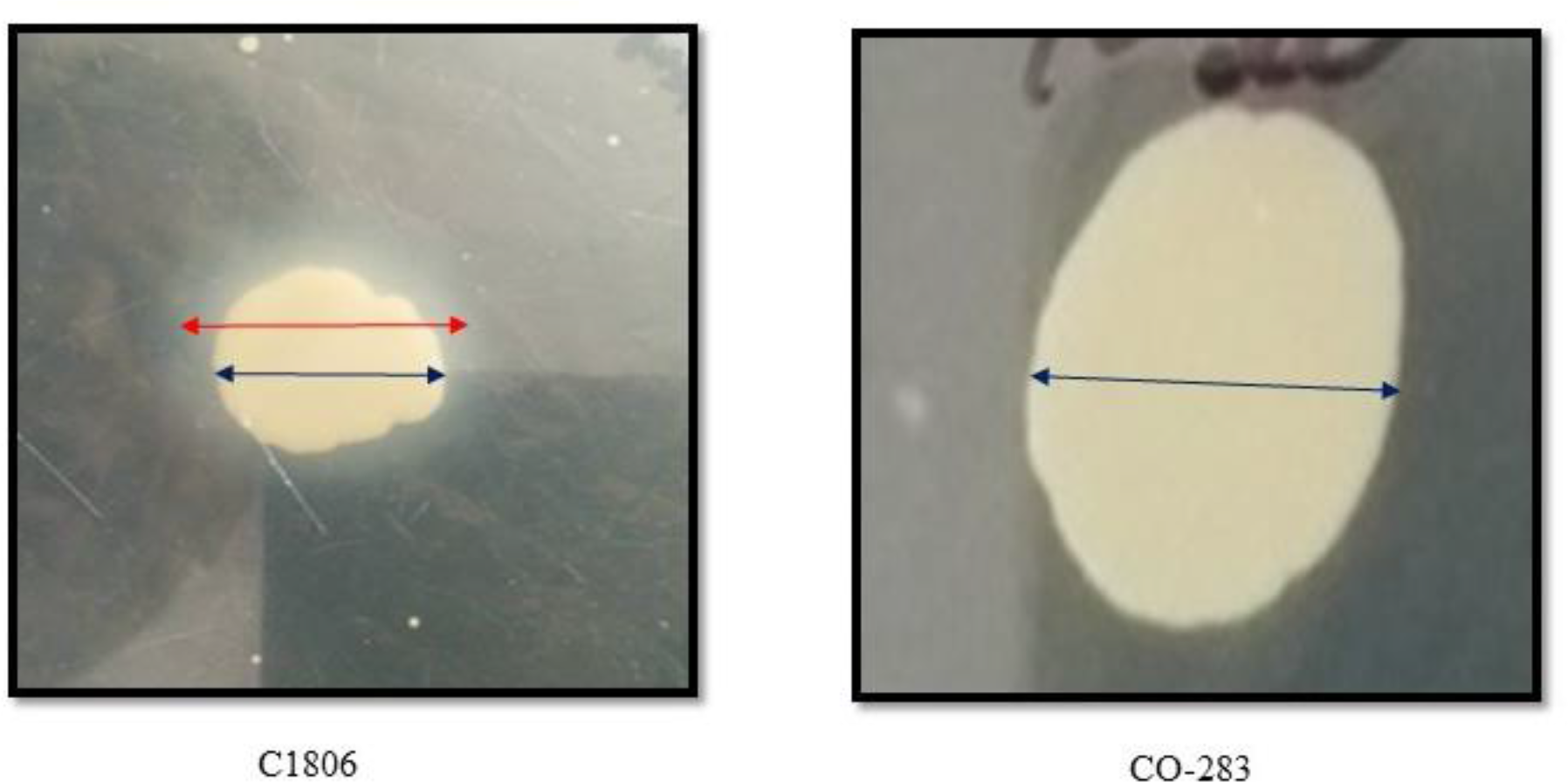
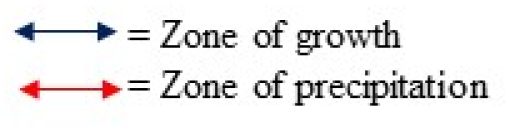
Phospholipase activity of the isolates: The figure shows the phospholipase activity of the isolates.

**Table S1:**
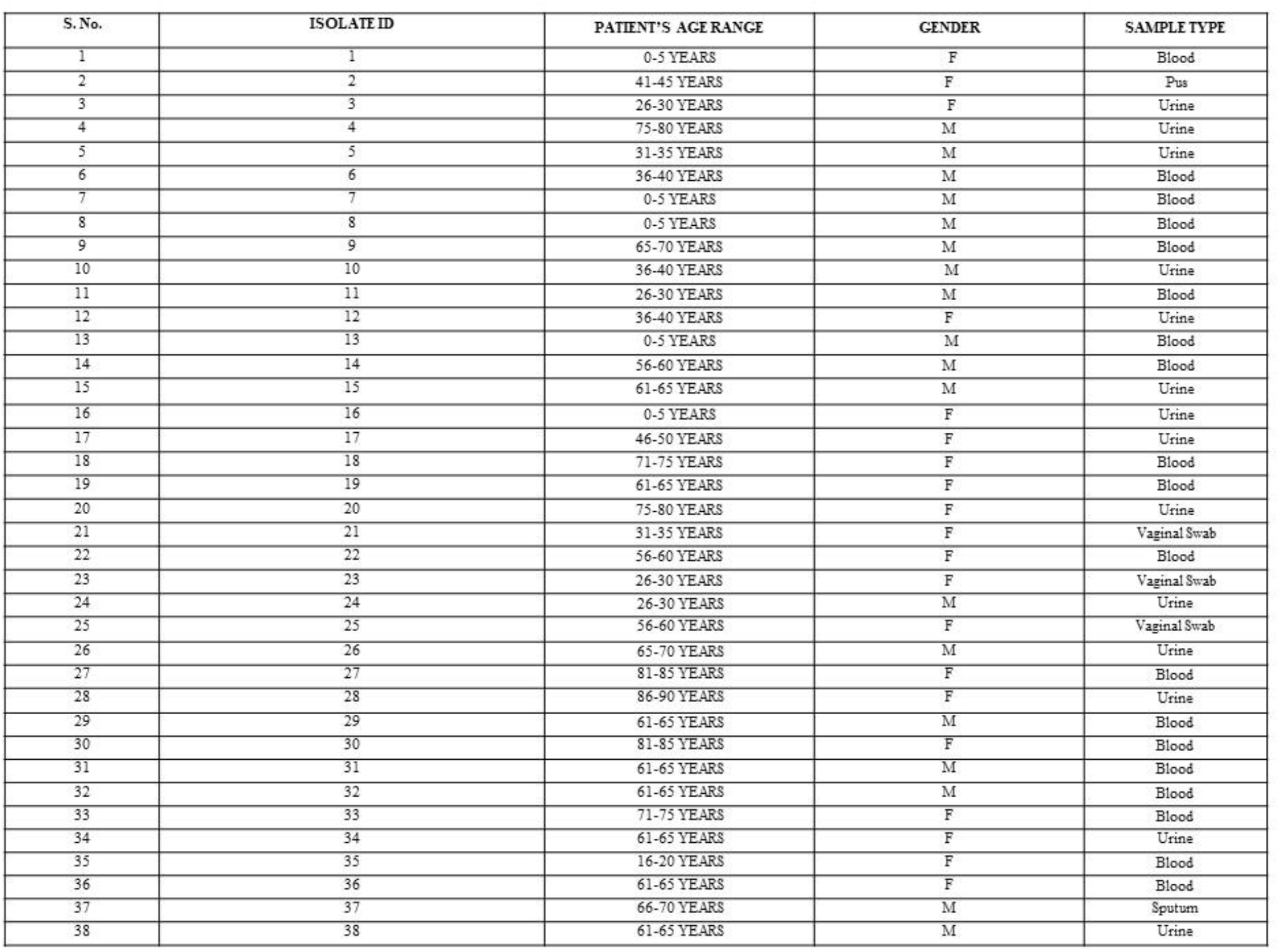
Demographic details of isolates: The table shows the age-range, gender and sample type of the isolates.

**Table S2:** Germ tube formation by the isolates: The table shows the germ tube formation capacity of the all isolates.

| S. No. | ISOLATE ID | NO. OF GERM TUBE POSITIVE | NO. OF TOTAL CELL COUNT | TOTAL RESULT (%) | SPECIES (A/NA) |
| --- | --- | --- | --- | --- | --- |
| 1 | 1 | 14 | 1012 | 1.3% | Albicans |
| 2 | 2 | - | - | Absent | Non-Albicans |
| 3 | 3 | 345 | 804 | 42.9% | Albicans |
| 4 | 4 | - | - | Absent | Non-Albicans |
| 5 | 5 | 15 | 1014 | 1.47% | Albicans |
| 6 | 6 | - | - | Absent | Non-Albicans |
| 7 | 7 | 24 | 1051 | 2.28% | Albicans |
| 8 | 8 | - | - | Absent | Non-Albicans |
| 9 | 9 | 8 | 1000 | 0.8% | Albicans |
| 10 | 10 | 14 | 1203 | 1.16% | Albicans |
| 11 | 11 | 52 | 1013 | 5.13% | Albicans |
| 12 | 12 | 121 | 1047 | 11.55% | Albicans |
| 13 | 13 | - | - | Absent | Non-Albicans |
| 14 | 14 | - | - | Absent | Non-Albicans |
| 15 | 15 | 32 | 1041 | 3.07% | Albicans |
| 16 | 16 | 80 | 1019 | 7.85% | Albicans |
| 17 | 17 | 207 | 1048 | 19.75% | Albicans |
| 18 | 18 | - | - | Absent | Non-Albicans |
| 19 | 19 | - | - | Absent | Non-Albicans |
| 20 | 20 | 5 | 1000 | 0.5% | Albicans |
| 21 | 21 | 6 | 1000 | 0.6% | Albicans |
| 22 | 22 | 12 | 1000 | 1.2% | Albicans |
| 23 | 23 | - | - | Absent | Non-Albicans |
| 24 | 24 | 21 | 1008 | 2.08% | Albicans |
| 25 | 25 | 28 | 1011 | 2.76% | Albicans |
| 26 | 26 | 46 | 1129 | 4.07% | Albicans |
| 27 | 27 | 25 | 1000 | 2.5% | Albicans |
| 28 | 28 | 15 | 1000 | 1.5% | Albicans |
| 29 | 29 | 39 | 1074 | 3.63% | Albicans |
| 30 | 30 | 16 | 1132 | 1.41% | Albicans |
| 31 | 31 | - | 1106 | Absent | Non-Albicans |
| 32 | 32 | 32 | 1107 | 2.89% | Albicans |
| 33 | 33 | - | 1102 | Absent | Non-Albicans |
| 34 | 34 | 16 | 1106 | 1.44% | Albicans |
| 35 | 35 | - | 1023 | Absent | Non-Albicans |
| 36 | 36 | - | 1054 | Absent | Non-Albicans |
| 37 | 37 | 215 | 1008 | 21.32% | Albicans |
| 38 | 38 | 18 | 1214 | 1.48% | Albicans |

**Table S3:** Comparative analysis of virulence factors: The table shows comparative analysis of the isolates between germ tube formation and phospholipase activity (Pz value).

| S.No. | ISOLATES ID | Germ Tube formation (%) | Pz VALUE |
| --- | --- | --- | --- |
| 1 | 1 | 1.3% | 0.34 |
| 2 | 2 | Absent | 1 |
| 3 | 3 | 42.9% | 0.3 |
| 4 | 4 | Absent | 0.71 |
| 5 | 5 | 1.47% | 0.4 |
| 6 | 6 | Absent | 1 |
| 7 | 7 | 2.28% | 0.37 |
| 8 | 8 | Absent | 0.57 |
| 9 | 9 | 0.8% | 0.75 |
| 10 | 10 | 1.16% | 0.51 |
| 11 | 11 | 5.13% | 0.75 |
| 12 | 12 | 11.55% | 1 |
| 13 | 13 | Absent | 0.67 |
| 14 | 14 | Absent | 1.07 |
| 15 | 15 | 3.07% | 0.89 |
| 16 | 16 | 7.85% | 1 |
| 17 | 17 | 19.75% | 0.76 |
| 18 | 18 | Absent | 0.85 |
| 19 | 19 | Absent | 1 |
| 20 | 20 | 0.5% | 0.85 |
| 21 | 21 | 0.6% | 0.84 |
| 22 | 22 | 1.2% | 0.78 |
| 23 | 23 | Absent | 1 |
| 24 | 24 | 2.08% | 0.95 |
| 25 | 25 | 2.76% | 0.75 |
| 26 | 26 | 4.07% | 0.42 |
| 27 | 27 | 2.5% | 0.73 |
| 28 | 28 | 1.5% | 1 |
| 29 | 29 | 3.63% | 0.89 |
| 30 | 30 | 1.41% | 0.79 |
| 31 | 31 | Absent | 1 |
| 32 | 32 | 2.89% | 0.80 |
| 33 | 33 | Absent | 1 |
| 34 | 34 | 1.44% | 0.76 |
| 35 | 35 | Absent | 0.80 |
| 36 | 36 | Absent | 1 |
| 37 | 37 | 21.32% | 0.49 |
| 38 | 38 | 1.48% | 0.40 |

## References

[1] Rafat, Z., Hashemi, S. J., Ahamdikia, K., Ghazvini, R. D., & Bazvandi, F. (2017). Study of skin and nail Candida species as a normal flora based on age groups in healthy persons in Tehran-Iran. Journal de Mycologie Médicale, 27(4), 501-505.

[2] Papon, N., Courdavault, V., Clastre, M., & Bennett, R. J. (2013). Emerging and emerged pathogenic Candida species: beyond the Candida albicans paradigm. PLoSpathogens, 9(9), e1003550.

[3] Bongomin, F., Gago, S., Oladele, R. O., & Denning, D. W. (2017). Global and multinational prevalence of fungal diseases—estimate precision. Journal of fungi, 3(4), 57.

[4] Arora, D. R., Saini, S., & Gupta, N. (2003). Evaluation of germ tube test in various media. Indian journal of pathology & microbiology, 46(1), 124-126.

[5] Fule, S. R., Das, D., & Fule, R. P. (2015). Detection of phospholipase activity of Candida albicans and non albicans isolated from women of reproductive age with vulvovaginal candidiasis in rural area. Indian Journal of Medical Microbiology, 33(1), 92-95.

[6] Tortorano, A. M., Rigoni, A. L., Biraghi, E., Prigitano, A., & Viviani, M. A. (2003). The European Confederation of Medical Mycology ECMM) survey of candidaemia in Italy: antifungal susceptibility patterns of 261 non-albicans Candida isolates from blood. Journal of Antimicrobial Chemotherapy, 52(4), 679-682.

[7] Sobel, J. D. (1989). Pathogenesis of Candida vulvovaginitis. Current topics in medical mycology, 86-108.

[8] Ghannoum, M. A. (2000). Potential role of phospholipases in virulence and fungal pathogenesis. Clinical microbiology reviews, 13(1), 122-143.

